# Assessing the effectiveness of multi-session online emotion recognition training in autistic adults

**DOI:** 10.1101/2024.07.23.24310558

**Authors:** Zoe E Reed, Oliver Bastiani, Andy Eastwood, Ian S Penton-Voak, Christopher Jarrold, Marcus R Munafò, Angela S Attwood

## Abstract

**Purpose:** Difficulties with emotion recognition can occur in neurodevelopmental conditions, including in autistic individuals. Providing interventions to support this would therefore be beneficial, particularly in terms of downstream effects on wellbeing, social relationships and education.

**Methods:** In this online experimental study, we examined the effect of a recently developed facial emotion recognition training task versus a sham/control task in an adult population identifying as autistic over four sessions in a 2-week period, with a fifth follow-up session (N=184).

**Results:** Our main analyses showed that facial emotion recognition accuracy was greater in Session 4 in the active group, with an estimated improvement of 14% (equivalent to approximately 7 additional correct responses), compared to 2% (equivalent to approximately 1 additional correct responses) in the sham group. Additional analyses suggested training effects were generalisable to facial stimuli that participants had not been trained on and were still present, although attenuated, two weeks later. We also observed some self-reported improvements in social interactions post-training.

**Conclusion:** Overall, this study demonstrated improved emotion recognition in an adult autistic sample with this training task. Future work is needed to investigate the effect of this emotion recognition training on emotion recognition accuracy in autistic children, where support could be most beneficial.

## Introduction

Difficulties with emotion recognition (ER) - the ability to recognise other’s emotional expressions - occur across a range of neurodevelopmental conditions, particularly in autistic individuals or those with autistic traits – those scoring higher on autistic trait questionnaires (Leung et al., 2022; Lozier et al., 2014; Uljarevic and Hamilton, 2013; Yeung, 2022). Studies have demonstrated that there are *global* (rather than emotion-specific) difficulties across all emotions in autistic individuals (Yeung, 2022). ER difficulties can negatively influence wellbeing and social skills, which in turn may negatively impact social relationships and educational outcomes such as school attendance (Adams, 2022; Kirst et al., 2022; Rice et al., 2015; Silveira-Zaldivar et al., 2020). Therefore, providing individuals with an intervention to help with ER is important and may have positive downstream benefits.

Effectiveness of a computer based-ER training task on emotion recognition accuracy in the general adult population has previously been demonstrated and it has been shown that the effects of this training transfer to facial stimuli other than those individuals were trained on (Reed et al., 2023). This task presents facial emotional expressions, of varying intensities, and asks the individual to select the emotion they believe was presented. They are then given feedback as to whether this was correct or not and if incorrect, they try again until a correct response is given. However, it is important to determine whether similar training effects are observed in autistic individuals who may experience greater difficulties in this area and who would be the key user-group for interventions.

This online experimental study therefore examined the effect of an ER training task versus a sham/control task on ER in an adult autistic population (or those identifying as autistic). Previous work examined training during a single session; however, it is unclear whether training over multiple sessions may be of additional benefit. Therefore, this study comprised four sessions of training over a 2-week period. We hypothesised that participants randomised to ER training would show greater improvement in ER ability, after the 4 sessions, compared to those randomised to sham training. We also explored whether: 1) ER training effects transferred to other (untrained) facial stimuli; and 2) there was evidence of continued ER improvement and impact on self-reported social interaction/skills two weeks after training completion.

## Methods

The protocol for this study was pre-registered on the Open Science Framework (https://osf.io/jszw7). Participants were recruited via the online recruitment platform Prolific (https://www.prolific.co/) and data collected via Gorilla, the online experiment builder (http://www.gorilla.sc/) (Anwyl-Irvine et al., 2019).

Data and code availability: The data and analysis code that form the basis of the results presented here for all studies are available from the University of Bristol’s Research Data Repository (http://data.bris.ac.uk/data/), DOI: To be added when published).

Compliance with Ethical Standards: This study received ethics approval from the School of Psychological Science Research Ethics Committee at the University of Bristol (approval code: 260821118826).

### Participants

A total of 220 participants were recruited and randomised to one of two training groups (active or sham) in a 1:1 ratio. To be eligible, participants needed to be autistic (self-reported diagnosis of autism) or identify as autistic (self-reported on Prolific), be aged 18 years or over, and be fluent in English. In addition, they could not: be currently taking medication to treat a mental health condition or medication usually prescribed for this; have an uncorrected visual impairment, including colour vision deficiency; have participated in any related studies (https://osf.io/x4kh3, https://osf.io/drby2 and https://osf.io/bpzcj); or participated in fewer than 10 studies on Prolific (to identify Prolific users more likely to complete all 5 sessions). All screening questions were self-reported by participants in their Prolific profiles, with further confirmation from them in Gorilla to verify eligibility. The exact screening questions used in Prolific are provided in Supplementary Materials Section 1. There were no restrictions based on geographical location.

Sample size was guided by a previous study investigating effectiveness of an emotional bias retraining task that uses the same stimulus set as the ER task tested here (Penton-Voak et al., 2012). An effect size of *d*=1.08 in the balance point (i.e., bias score in a single emotion training study) was reported in that work. A more conservative effect size of *d*=0.50 was used in our sample size calculation to account for the current task training six emotions simultaneously (compared to two in the bias version of the task) (Ioannidis, 2008), and an extra 5% was added to the sample size due to potential attrition. At an alpha level of 5% (i.e., p=0.05) for a two-tailed independent means t-test one would need 210 participants to provide 95% power to detect an effect size of *d*=0.50. Therefore, we recruited a total of 220 participants (accounting for the extra 5% needed).

### Study procedure

The study consisted of a total of 5 sessions to assess the effect of ER training over time, with 4 of these being training sessions and the primary outcome (total number of correct responses on the ER task) assessed after training in session 4. The 5 sessions were completed over a 3– 4-week period, with each of the first 4 sessions completed at least 24 hours after the previous session and Session 5 completed approximately 2 weeks after Session 4 (see Fig. 1). If any participants did not complete session 2 by the end of day 12, they were not invited back for session 3 and they were replaced. In addition, if a participant did not complete all 4 sessions they were replaced.

**Fig. 1.**
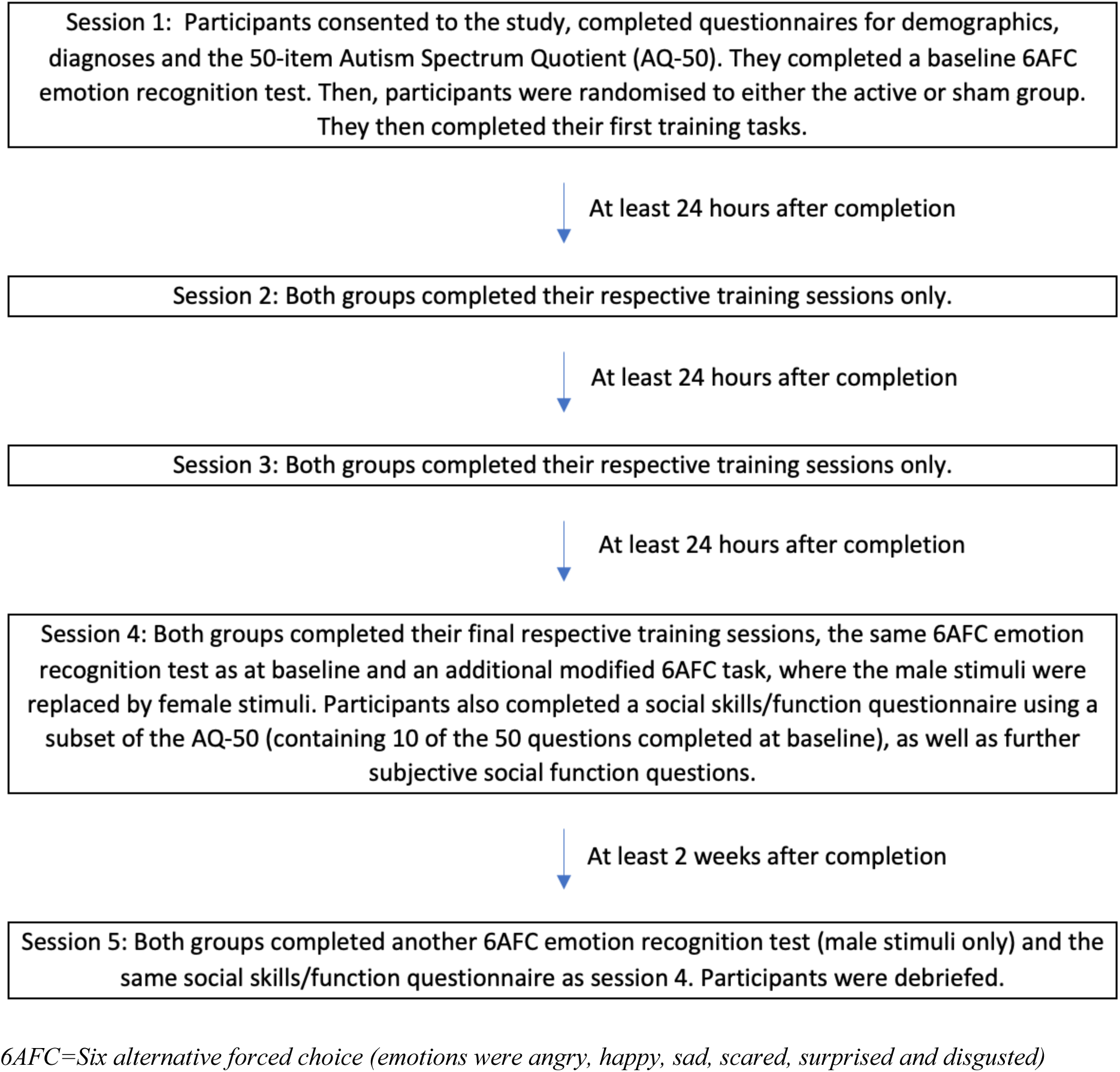
Study session overview.

### Demographic information

Demographic information on age, gender and education were collected in Session 1. For gender, participants were asked ‘What gender do you identify as?’ and they could select from male, female, and non-binary. For education, participants were asked ‘What is the highest level of education you have completed?’ and they could select from ‘Degree or equivalent higher education and above’, ‘A level or equivalent’ (A levels: these are UK subject specific qualifications that are typically completed over 2 years between the ages of 16 and 18), ‘GCSEs grades A*-C or equivalent’ (General Certificate of Secondary Education: UK subject-specific qualifications typically completed over 3 years towards the end of secondary school education), ‘No qualification’, or ‘I don’t know’. The last two options were combined for analysis.

### Emotion recognition test – Six Alternate Forced Choice (6AFC)

The ER test was included in Sessions 1 (baseline), 4 (primary outcome) and 5 (follow up – secondary outcome). On each trial, participants were presented with a single facial image (the same white male face for all). These stimuli were computer-generated by averaging photos of 12 individuals, and therefore do not show an identifiable person (Dalili et al., 2016). Each stimulus expressed one of six emotions (happy, angry, sad, scared, surprised, and disgusted) at one of 8 levels of intensity (neutral to 100% of that emotional expression). Therefore, there were a total of 48 trials, which were each displayed once and were shown in a random order. Facial emotion expression images were presented on screen for 150 milliseconds (ms), before being masked for a further 250ms, and then participants proceeded to the next screen with the six emotions displayed as words. Here, participants were asked to select the word that they thought represented the displayed emotion, with the selection screen remaining present until participants had made their choice. After the choice was made no feedback was provided and participants moved onto the next image, preceded by a fixation cross. Further details of this test can be found in elsewhere (Reed et al., 2023).

### Generalisability test

In Session 4, post-training, participants completed a further ER test (with the same parameters as the main ER test), but with white female facial stimuli instead. The purpose of this was to test whether effects of training would generalise to non-trained faces.

### Emotion recognition training task

The active training group completed a modified version of the ER test, whereby the procedure remained the same, with exceptions that each face was displayed for 1000ms, and participants selected an emotion word until they were correct. Feedback was presented to participants after each selection, and they could only proceed to the next face image once they had answered correctly. Fig. 2 shows an example of the ER test task and further details can be found elsewhere (Reed et al., 2023).

**Fig. 2.**
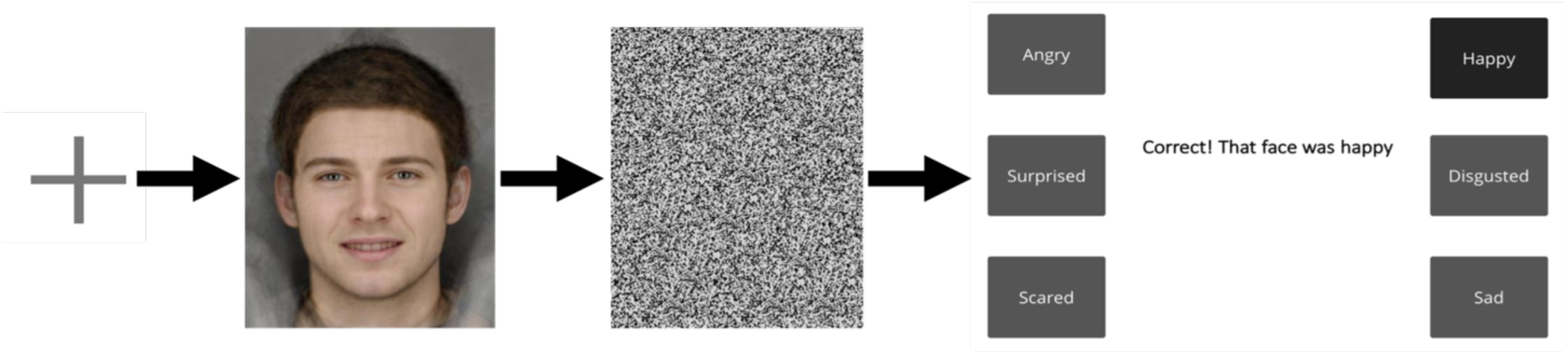
Example of ER test task

### Sham training task

For the sham training group, a similar training task was completed, however instead of faces with emotional expressions, the stimulus images presented were of coloured rectangles (blue, red, green, yellow, purple and orange, all ranging from grey to the full colour with 8 increments), with participants being asked to select which colour they had seen displayed on the preceding screen. They were also provided with feedback, and they could only proceed to the next image once they had answered correctly.

### Emotion recognition outcome measures

The primary outcome for statistical analyses was the total number of correct responses i.e., the number of times participants correctly selected the emotion corresponding with the facial expression displayed (hits). This was used as an indicator of ER accuracy in the baseline and post-training (Session 4) tests.

Outcomes for secondary analyses were 1) total hits from the post-training generalisability test, 2) total hits in Session 5, approximately 2 weeks post-training (i.e., to examine effects after a time delay), 3) emotion-specific sensitivity scores (using the signal detection parameter A-prime (A’) index which is a non-parametric estimate of discriminability) (Pallier, 2002) post-training in Session 4 (analyses were also conducted for hits and false alarms – the total number of times an emotion was selected when this was not the correct response – which are factored into the sensitivity scores and which are presented in the Supplementary Materials). Hits and false alarm outcomes across all analyses were converted to proportions.

### Autism and mental health diagnoses

As not all participants recruited had a diagnosis of autism (some identified as autistic or were going through the diagnosis process), participants were asked whether they had ever received a formal diagnosis of autism to allow us to explore whether this had an impact on the results. Participants were also asked whether they had a diagnosis of anxiety, depression, or other mental health issue to assess whether co-occurring mental health conditions impacted results.

### Autistic traits

The full 50-item Autism Spectrum Quotient (AQ-50) questionnaire was included to measure autistic traits (Baron-Cohen et al., 2001). The total possible score was 50, with higher scores indicating more autistic traits. For each statement in the AQ-50, participants were asked to choose the response that best describes them from the following options: ‘Definitely Agree’, ‘Slightly Agree’, ‘Slightly Disagree’ or ‘Definitely Disagree’. The full AQ-50 is provided in Supplementary Materials Section 2.

A 10-item subset of the Autism Spectrum Quotient (see Supplementary Materials Section 2) was used to measure social skills specifically and data from this were collected in Sessions 4 and 5 (as well as part of the full AQ-50 in Session 1). The score on this subset of questions was used to assess whether there were any changes in social skills following training.

### Other social skills questions

In Sessions 4 and 5, additional questions were asked to measure any subjective changes in social skills not picked up by the social skills subset of the AQ-50. Participants were asked ‘Has the frequency of your social interactions increased since participating in this study?’ and ‘Do you feel that this study has improved your ability to recognise other people’s emotions?’. These were rated on a scale of 0-100 where 0 indicated “not at all” and 100 indicated “very much so”.

### Subjective ratings of training

To assess whether there were group differences in how they perceived the study participants were asked whether they found the tasks tiring and interesting and whether the instructions were easy to follow. They were also asked whether they thought the ER training would be useful for autistic individuals. The exact questions asked are provided in Supplementary Materials Section 3. These were rated on a scale of 0-100 where 0 indicated “not at all” and 100 indicated “very much so”.

## Statistical analysis

### Primary analysis

All analyses were conducted in R version 4.0.2 (R Core Team, 2016). Prior to analysis we removed individuals whose total hits scores were outliers in the baseline and Session 4 post-training tests (i.e., data points that fell 1.5 times above or below the interquartile range). Then data were also assessed for normality using skewness and kurtosis statistics. A linear mixed effects (LME) model compared group differences (active versus sham training) for total hits (the number of correct responses), accounting for between participants random variance, with variables of time (baseline and Session 4 – the primary outcome), group, and an interaction term for time x group. This was conducted using the lme4 package in R (Bates et al., 2015). Random intercepts for each participant ID were included for the random effects. We ran the following models: 1) an unadjusted model, 2) a model adjusted for age, gender and education level (as fixed effects), and 3) a model additionally adjusted for scores on the AQ50 at baseline.

### Secondary analyses

We conducted several secondary analyses to examine generalisability to non-trained stimuli, maintenance of effects over time, emotion-specific effects and wider impacts of ER training (i.e., on social skills and subjective ratings of the training).

First, we ran a similar model to our primary analysis but the outcome for Session 4 was hits on the generalisability test.

Second, we examined whether any training effects were maintained after approximately 2 weeks by running a similar LME model to that for the primary analysis but instead using hits data from Session 5 instead of Session 4.

Third, we ran analyses to explore sensitivity scores across the individual emotions (as well as hits and false alarm rates). This was achieved by running LME models for each emotion for sensitivity scores (or hits or false alarms) as outcomes with the same variables as in the adjusted primary outcome model.

Fourth, to assess whether there was any transfer of ER training effects onto social skills we also ran similar LME models to those in the primary analysis (adjusted), but instead using the AQ-50 social skills subset as the outcome for Session 4 and for Session 5, as two separate models.

Fifth, exploration of the data from the other subjective social skills questions asked in Sessions 4 and 5 was conducted using t-tests to compare the groups. We also assessed whether each of the subjective ratings of training experiences (i.e., sham vs active) varied between the two groups by conducting two-tailed independent means t-tests.

Other pre-registered exploratory analyses are provided in the Supplementary Materials Section 4.

### Sensitivity analyses

We conducted additional sensitivity analyses for our primary analyses and secondary analyses with hits in Session 5 by 1) excluding participants with any other mental health diagnosis to see whether having a co-occurring mental health diagnosis impacted total hits and 2) excluding participants who had encountered technical issues during the study (e.g., completing a session over multiple days or 2 on the same day, which could have impacted their outcome).

## Results

### Participant characteristics

A total of 220 participants were recruited in Prolific. However, not all participants met eligibility criteria when asked the screening questions in Gorilla (N=20) and therefore these individuals were excluded from analyses (post-randomisation). In addition, we excluded participants who did not have complete data for Session 4 (N=9) or had outliers in their data for total hits at baseline or in Session 4 (N=7). Therefore, 184 individuals were included in the analyses (94 in the active and 90 in the sham groups). Table 1 shows a description of the sample. After removing outliers, we examined skewness and kurtosis for baseline and post-training (Session 4) total hits. Histograms of these distributions are shown in Supplementary Fig. S1. Skewness and kurtosis measures were within an acceptable range (see Supplementary Materials Section 5 for further details).

**Table 1.**
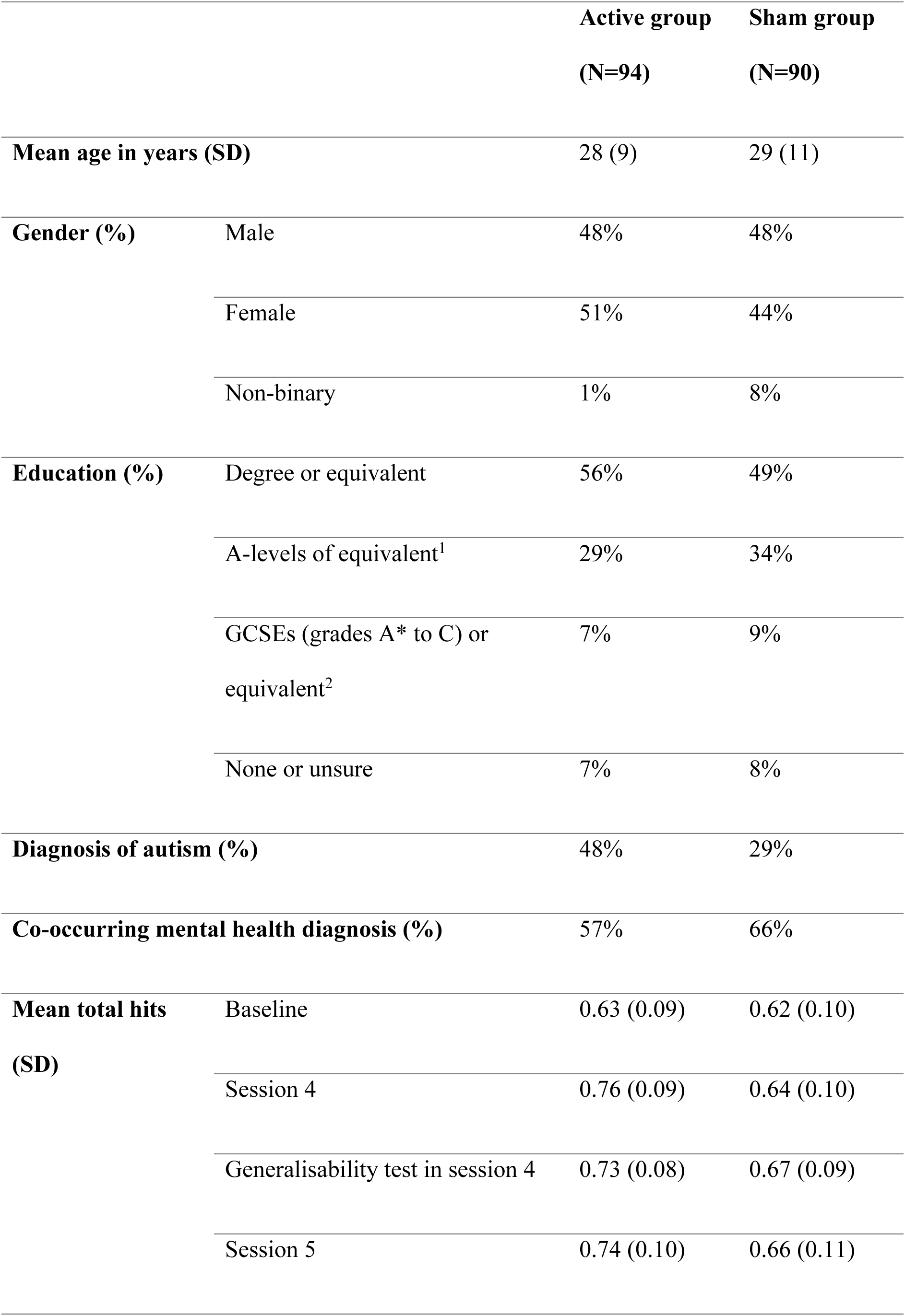
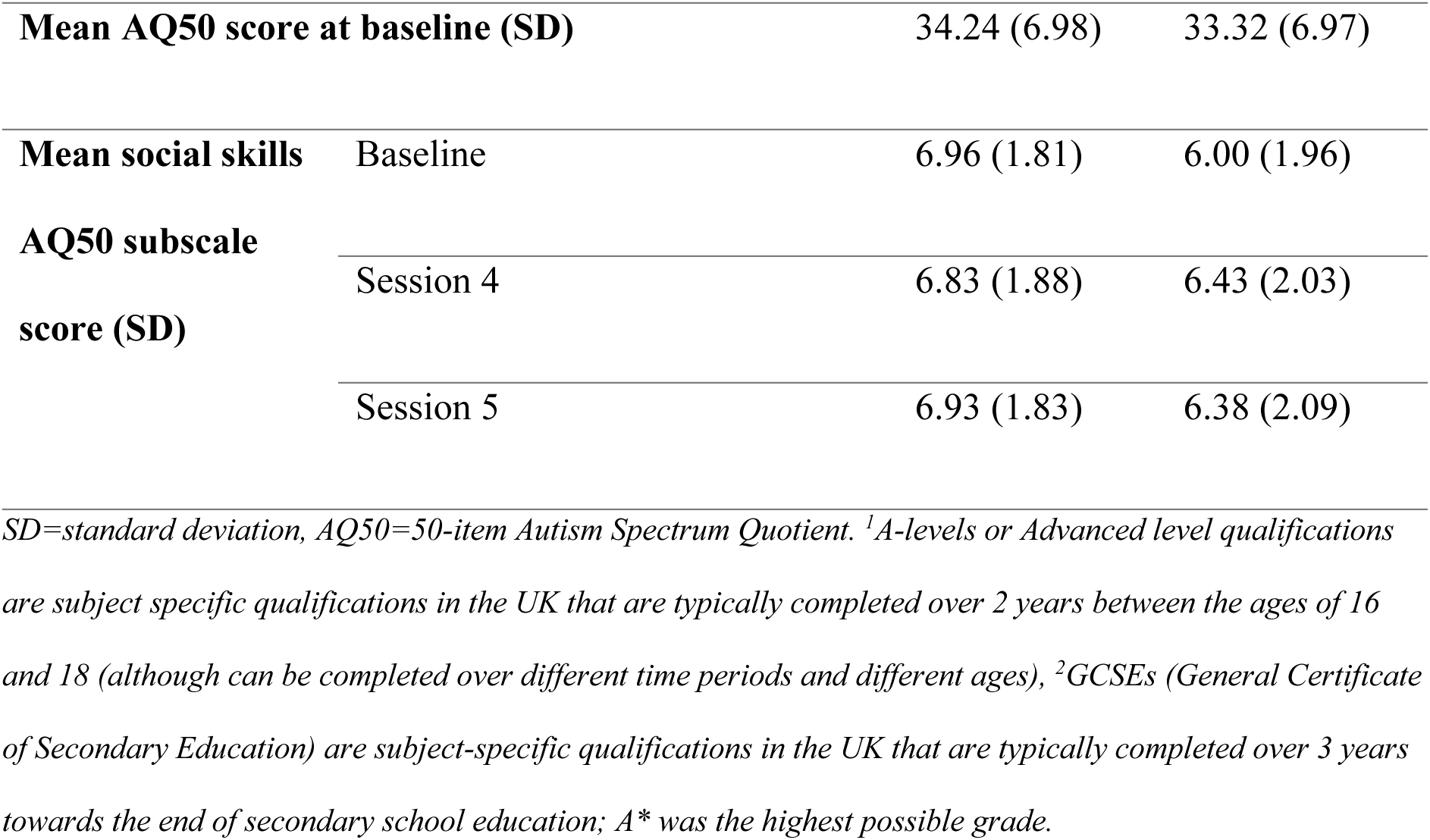
Participant characteristics.

### Primary analysis results: emotion recognition accuracy in Session 4

Analyses indicated that the active group showed greater improvement post-training in Session 4 compared to the sham group. Specifically, the interaction model revealed that the proportion of total hits was greater in Session 4 in the active group compared to the sham group (see Fig. 3 and Supplementary Table S1) in the unadjusted model and the model including age, gender and education level (including covariates: *b*=0.12, 95% CI=0.08 to 0.16, *p*=4x10^-09^). In the fully adjusted model, the sham group hits increased from an estimated 67% at baseline to 69% in Session 4, whilst the active group increased from an estimated 67% to 81%. Results were similar in the model that additionally included scores on the AQ50 at baseline (see Supplementary Table S1).

**Fig. 3.**
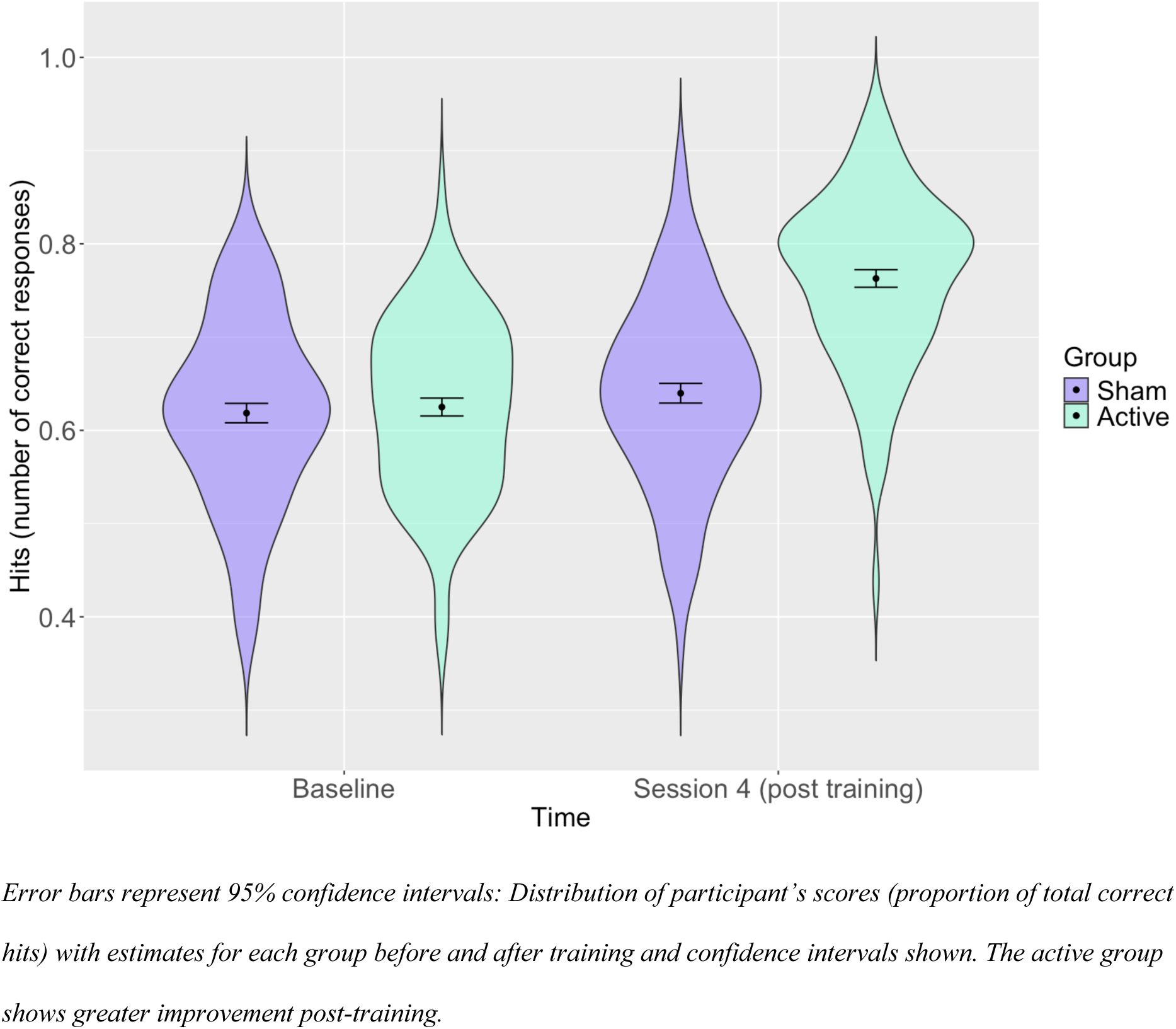
Distribution of participants’ total hits with estimates for the active and sham groups at baseline and post training (Session 4).

### Secondary analysis results

#### Generalisability

In the generalisability test (Fig. 4 and Supplementary Table S3) we found slightly attenuated results but there was still a clear indication of greater hits post-training in the active group than the sham group (*b*=0.06, 95% CI=0.02 to 0.09, *p*=0.005), with the sham and active group’s hit count increasing from an estimated 67% to 72% and 78%, respectively.

**Fig. 4.**
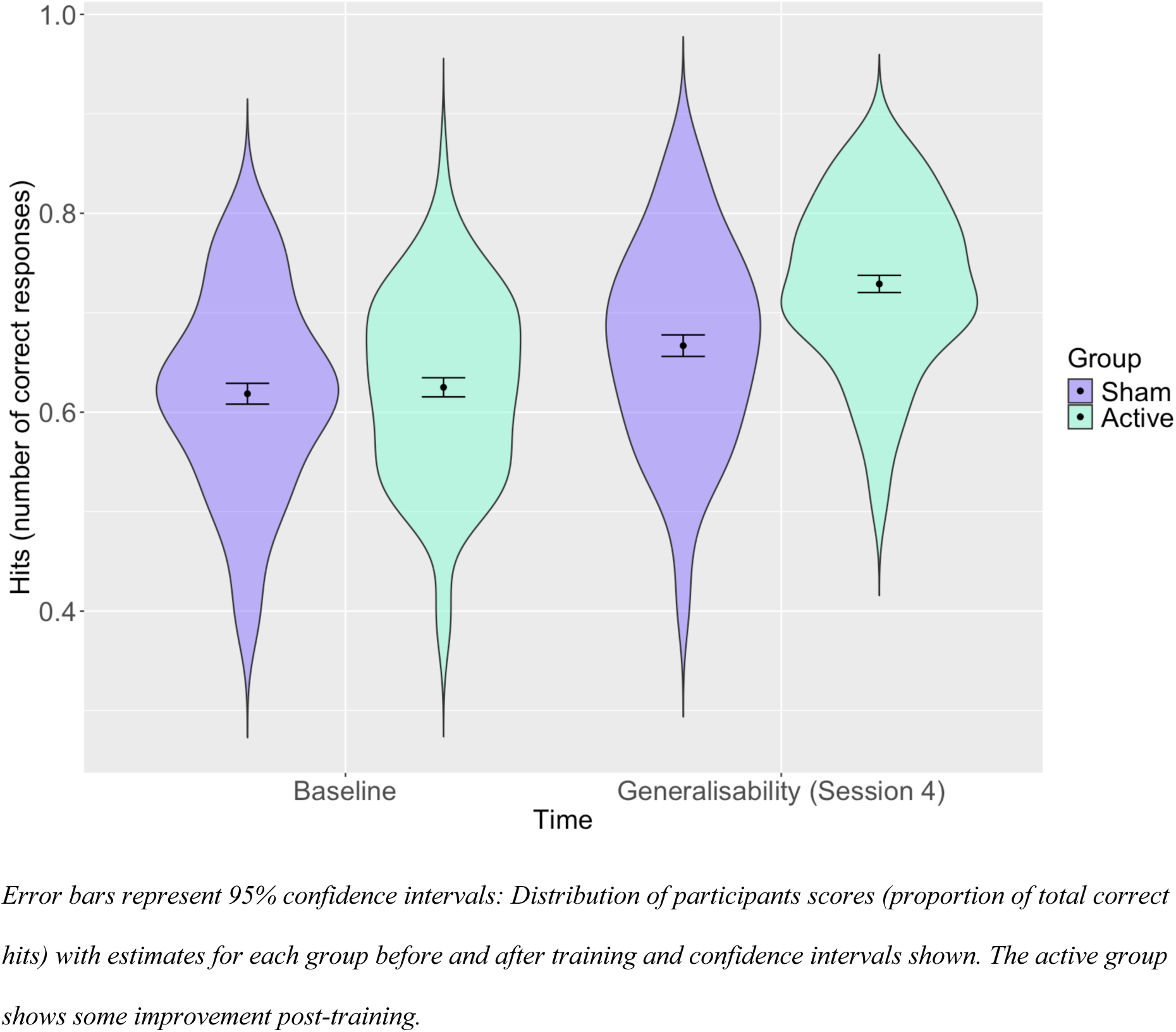
Distribution of participants’ total hits with estimates for the active and sham groups at baseline and in the post training generalisability test.

#### Maintenance

There was some attenuation of the training effect at Session 5 (2 weeks post-training) for the active group (Fig. 5 and Supplementary Table S4), but this still remained, indicating that this selective improvement persisted over time (*b*=0.07, 95% CI=0.03 to 0.11, *p*=0.001), with the sham and active group’s hit count increasing from an estimated 67% and 68% to 71% and 79%, respectively.

**Fig. 5.**
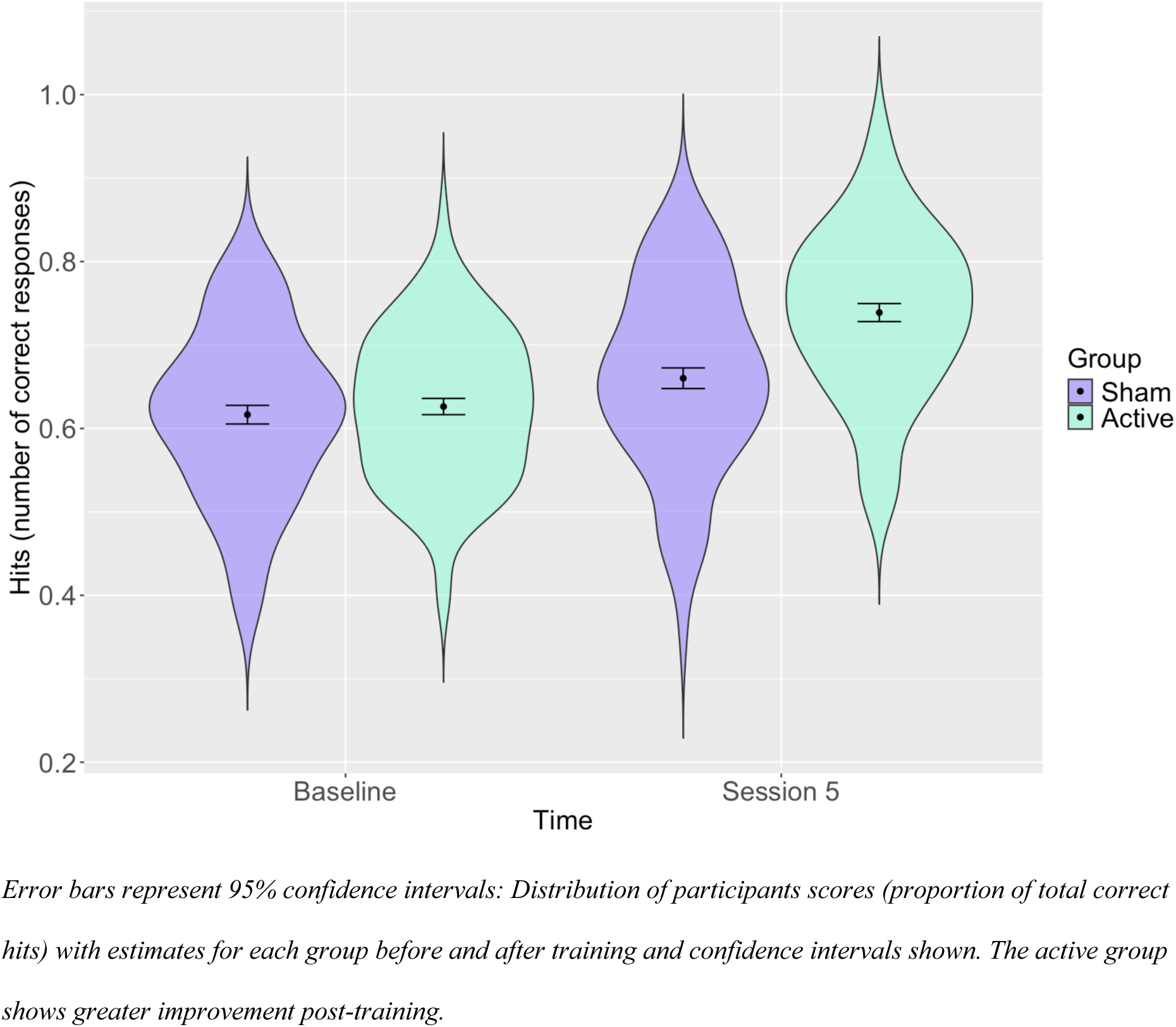
Distribution of participants’ total hits with estimates for the active and sham groups at baseline and 2 weeks post training (Session 5).

#### Emotion specific models

Results from the LME models examining emotion-specific sensitivity scores, hits and false alarm rates are presented in Supplementary Tables S5 to S7. For sensitivity scores the was evidence of interaction effects between time and group for all emotions except surprised and disgust, where the active group showed higher scores (indicating greater discriminability) compared to the sham group post-training. For hits there was evidence of interaction effects between time and group for all emotions except disgust. This indicated that the active group recognised all emotions except disgust better than the sham group post-training. Finally, for false alarms there was evidence of interaction effects between time and group for the emotions of scared, surprised and disgusted, where the active group had fewer false alarms for these emotions compared to the sham group post-training.

#### Social skills

The results from LME models examining whether there was any transference of ER training effects onto social skills (as measured using a subset of the AQ-50) indicated that there was no meaningful difference in the active group compared to the sham group post-training after Session 4 (*b*=0.04, 95% CI=-0.31 to 0.39, *p*=0.83) (Fig. 6a), or Session 5 (*b*=0.32, 95% CI=-0.01 to 0.66, *p*=0.06) (Fig. 6b and Supplementary Table S8).

**Fig. 6.**
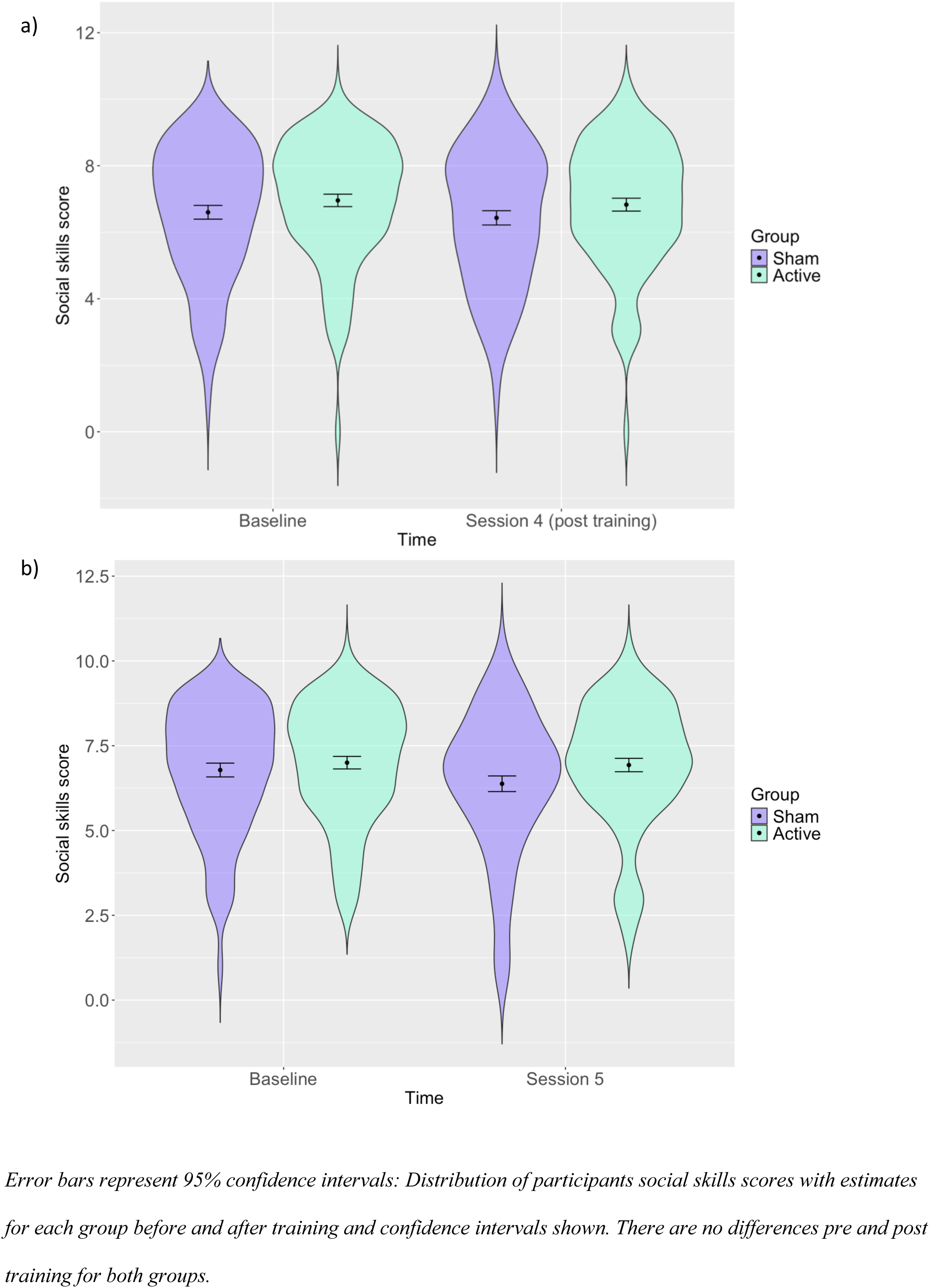
Distribution of social skills as measured by a subset of the AQ-50 at baseline and post-training in Session 4 (a) and Session 5 (b)

#### Subjective ratings

We found that there were some group differences in subjective ratings of the study (Supplementary Table S9). In particular, the active group found the study slightly more interesting (active mean=62 [SD=27], sham mean=52 [SD=25], *p*=0.01) and easier to follow (active mean=96 [SD=9], sham mean=90 [SD=15], *p*=0.002) than the sham group, but there were no meaningful differences observed for how tiring the study was (active mean=48 [SD=27], sham mean=45 [SD=29], *p*=0.51). In addition, differences were found between the groups for the question relating to ability to recognise other people’s emotions at the end of Session 4 (i.e., higher in the active group) (active mean=40 [SD=27], sham mean=26 [SD=23], *p*=0.0002) and this was rated even higher after Session 5 (active mean=47 [SD=24], sham mean=33 [SD=22], *p*=9.19x10^-05^), although we note that values were not very high in the overall measure out of 100. Additionally, although meaningful group differences in self-reported frequency of social interactions after Session 4 were not observed (active mean=20 [SD=26], sham mean=17 [SD=33], *p*=0.40), a meaningfully greater frequency of social interactions was seen after Session 5 in the active compared to the sham group (active mean=38 [SD=30], sham mean=28 [SD=28], *p*=0.03). Finally, there was no meaningful difference in the active and sham groups for ratings of how useful ER training would be for autistic individuals, but we note that ratings were supportive overall of this training (active mean=69 [SD=23], sham mean=68 [SD=21], *p*=0.88).

### Sensitivity analysis results

Results were similar to the primary analyses with Session 4 hits as the outcome and secondary analyses with Session 5 hits as the outcome, when excluding participants with 1) any other mental health diagnosis and 2) who had encountered technical issues during the study (Supplementary Tables S2 and S4).

## Discussion

We examined whether active ER training improved ER of facial expressions compared with sham training in autistic adults. Our results indicate an estimated improvement of 14% (equivalent to approximately 7 additional correct responses) in the active group compared to 2% (equivalent to approximately 1 additional correct responses) in the sham group in our adjusted models, demonstrating effectiveness of ER training in autistic individuals. These results build on a previous study in a non-autistic sample (Reed et al., 2023). In the present study we observed a greater overall improvement in ER compared to a previous study which included only one session. This may indicate that a greater number of sessions is more beneficial, but, as the samples were different, further studies are needed to confirm this.

We also examined the generalisability of this ER training. Although our results were slightly attenuated in the generalisability test, we still observed an effect suggesting transference of training to novel facial stimuli, although further testing of this with a wider range of stimuli (including different ages and ethnicities) is needed in future work. Similarly, our results were attenuated, but with an effect still observed, approximately 2 weeks after the last training session indicating that there is a lasting effect of the training. It is unclear how long this effect may be sustained for and therefore further studies with longer follow-up periods would be useful.

The emotion-specific analyses revealed some group differences after training, particularly in the cases of disgust, scared, and surprised where the training had less of an effect than for the other emotions. There is evidence to suggest that specific emotions, in particular disgust, may be more difficult to recognise for autistic individuals (Law Smith et al., 2010). Thus, it may be that further and more tailored training is needed for these specific emotions, although it could also be the case that these expressions are particularly difficult to distinguish from one another.

We did not observe any meaningful differences in the social skills measure post-training, although this is not necessarily surprising given that the social skills measure is a subset of the autistic traits measure. We did observe some evidence of group differences post training on greater self-reported ER improvements and social interactions in the active group, which were even more apparent 2 weeks after the training.

Overall, our results are in line with previous studies demonstrating improved ER after training (Berggren et al., 2018; Farashi et al., 2022; Zhang et al., 2021). Our study also suggested effects are maintained two weeks post-training, whereas a previous meta-analysis suggested that effects were not maintained, although follow-up times were variable and not included in all studies, so their conclusions were limited (Zhang et al., 2021). Most of the previous studies in this area did not find evidence to suggest social skills improved post training, in line with our findings (Berggren et al., 2018; Zhang et al., 2021). However, our self-reported responses suggest there might be some improvement from a subjective perspective, suggesting that this should be examined in greater depth in future studies. There is limited information on generalisability in previous studies (Berggren et al., 2018; Zhang et al., 2021). Therefore, it is difficult to compare our generalisability results to previous studies, confirming that this is an area that requires further study. Our results suggest that this task would be useful to include in future research in this area.

### Limitations

Our study, whilst conducted in a well-powered sample over multiple sessions, is subject to some limitations. First, the stimuli used in all sessions were of the same individual; a decision taken to avoid making the task too long, particularly given its use over multiple sessions. As a result, although we found that effects were generalisable to other non-trained facial stimuli this needs to be tested for other facial stimuli (e.g., facial stimuli with different ages and ethnicities).

Second, we are unable to determine whether the effects observed are due to mere exposure effects (of faces) – individuals becoming more familiar with the facial stimuli as opposed to the training component of the task influencing emotional processing ability. Exposure effects are not necessarily problematic if the result is still ER being supported in the real world. In addition, previous studies using the same facial stimuli for bias retraining (which similarly has a feedback component) demonstrate that training effects transfer to untrained facial stimuli (Dalili et al., 2016; Griffiths et al., 2015). Third, this work similarly cannot distinguish the mechanisms behind any improvements in ER and it is important to consider that autistic individuals may have a different approach to emotion recognition than neurotypical individuals and that there will likely be differences within the autistic population, i.e., not all autistic individuals will experience ER difficulties.

Fourth, a limitation of our study is that we conducted the study online (due to the COVID-19 pandemic). This may impact our findings in several ways: i) we recruited from individuals signed up to Prolific which likely resulted in a selected sample, therefore future work would ideally be conducted by engaging individuals from across the community, ii) there is no way to really verify who has taken part in the study beyond Prolific’s checks or how well the participants engaged/paid attention to the study and iii) we cannot be sure how well these results generalise to real world settings (e.g., with children in a classroom), so this would need to be examined further. However, by conducting the study online we were able allow autistic individuals to take part in a more accessible way. Fifth, our results may be limited by the fact that a large proportion of the sample did not have a diagnosis of autism. We included those also identifying as autistic to be more inclusive in our research, however this may mean that those who would not meet diagnostic criteria are included. Finally, although we screened for colour vision deficiencies it may be that a participant is unaware that they have a colour vision deficiency, and therefore this may have impacted the sham task with colours. Given that this was a sham task this is unlikely to influence our results for ER, but alternative tasks could be considered in the future avoiding the use of colours.

### Future directions

This study demonstrated improvement in ER post training. Further studies would be useful to examine the extent of the training in more detail. For example, comparable studies with additional stimuli including different genders, ages and ethnicities would be useful to further explore generalisability. Future work should also examine differences in ER within autistic populations in order to create tasks that are tailorable to individuals as opposed to using a ‘one size fits all approach’, particularly through co-design with autistic individuals. In addition, in this study we worked with autistic adults, but future work with children would be useful to examine training further, as childhood is likely where most individuals would need support in this area, and where positive impacts on downstream outcomes would be more likely. Finally, further studies with other validated social interaction measures, which the autistic community consider to be useful to examine, would be beneficial in order to ascertain whether there are improvements in these areas. Both groups thought that ER training would be useful for autistic people, which is reassuring for future research in this area. Future research is needed to i) determine the optimal number of sessions, because in the current study, the number of hits continued to increase over sessions, and it is unclear how this would change over further sessions, ii) examine the downstream impact of ER training beyond just improving ER. Future research in this area should be conducted with input from the autistic community to create ER tasks which truly support autistic individuals who choose to receive support in this area.

### Conclusion

Overall, we found that multi-session ER training improved ER in an adult autistic sample. We additionally observed ER improvements that remained over time, and transferred to novel facial stimuli, and which may have a positive impact on social engagement and self-reported ER. Although further work is needed to determine: 1) whether one would see these improvements in autistic children, and 2) if there is transference to further stimuli and the real-world emotions, and whether improvements in ER have further downstream impacts, this study provides a good evidence base for this form of training task. It therefore provides a basis for further development of ER training tasks to support autistic individuals with ER difficulties.

## Data Availability

The data and analysis code that form the basis of the results presented here for all studies are available from the University of Bristol's Research Data Repository (http://data.bris.ac.uk/data/), DOI: To be added when published).

## Acknowledgements

We would like the thank all of the research participants that took part in our study. For the purpose of open access, the author(s) has applied a Creative Commons Attribution (CC BY) licence to any Author Accepted Manuscript version arising from this submission.

## Author contributions

Conceptualization: ZER, ASA; Methodology: ZER, AE, ASA; Formal Analysis: ZER; Investigation: ZER, OB; Resources: ZER, ASA; Data Curation: ZER; Writing—Original Draft: ZER, OB; Writing—Review and Editing: ZER, OB, AE, ISPV, CJ, MRM, ASA; Supervision: ASA, MRM; Project Administration: ZER, ASA; Funding Acquisition: ZER, MRM, ASA, ISPV.

