## Supplementary Materials for "Assessing the effectiveness of multi-session online emotion recognition training in autistic adults"

**1. Screening questions from Prolific**

Potential Prolific participants were only invited to take part based on responses to Prolific’s screening questions as detailed below. These were then rechecked in Gorilla.

“Have you received a formal clinical diagnosis of autism spectrum disorder, made by a psychiatrist, psychologist, or other qualified medical specialist? This includes Asperger’s syndrome, Autism Disorder, High Functioning Autism or Pervasive Developmental Disorder”.

We included individuals who selected yes (as a child or as an adult), who were in the process of receiving a diagnosis or who identified as being on the autism spectrum.

“What is your date of birth?”

We included individuals who were aged 18 years or over.

“Which of the following languages are you fluent in?”

We included individuals who selected English.

“Are you currently taking any medication to treat symptoms of depression, anxiety or low mood (e.g., SSRIs)?”

We excluded individuals who selected yes for any of the medications listed (anti-depressants, anti-anxiety medication, anti-psychotics).

“Do you have normal or corrected-to-normal vision?”.

We excluded participants who selected no.

We also excluded individuals who participated in any of related studies (<https://osf.io/x4kh3>, <https://osf.io/drby2> and <https://osf.io/bpzcj>); as they would be familiar with the task, and individuals who had participated in fewer than 10 studies on Prolific (to identify Prolific users more likely to complete all 5 sessions).

**2. Autism-spectrum quotient (AQ-50)**

Items included in the autism-spectrum quotient (AQ-50) (Baron-Cohen et al., 2001).

Participants will be asked to choose one response that best suits how each statement describes them using one of the following responses, “Definitely Agree”, “Slightly Agree”, “Slightly Disagree” or “Definitely Disagree”. For the following items a point is scored for the responses “Definitely Agree” or “Slightly Agree”: 1, 2, 4, 5, 6, 7, 9, 12, 13, 16, 18, 19, 20, 21, 22, 23, 26, 33, 35, 39, 41, 42, 43, 45, 46. For the remaining items, a point is scored for the responses “Definitely Disagree” or “Slightly Disagree”, these items are: 3, 8, 10, 11, 14, 15, 17, 24, 25, 27, 28, 29, 30, 31, 32, 34, 36, 37, 38, 40, 44, 47, 48, 49, 50. The 50 questions are:

1. “I prefer to do things with others rather than on my own.” *
2. “I prefer to do things the same way over and over again.”
3. “If I try to imagine something, I find it very easy to create a picture in my mind.”
4. “I frequently get so strongly absorbed in one thing that I lose sight of other things.”
5. “I often notice small sounds when others do not.”
6. “I usually notice car number plates or similar strings of information.”
7. “Other people frequently tell me that what I’ve said is impolite, even though I think it is polite.”
8. “When I’m reading a story, I can easily imagine what the characters might look like.”
9. “I am fascinated by dates.”
10. “In a social group, I can easily keep track of several different people’s conversations.”
11. “I find social situations easy.” *
12. “I tend to notice details that others do not.”
13. “I would rather go to a library than to a party.” *
14. “I find making up stories easy.”
15. “I find myself drawn more strongly to people than to things.” *
16. “I tend to have very strong interests, which I get upset about if I can’t pursue.”
17. “I enjoy social chitchat.”
18. “When I talk, it isn’t always easy for others to get a word in edgewise.”
19. “I am fascinated by numbers.”
20. “When I’m reading a story, I find it difficult to work out the characters’ intentions.”
21. “I don’t particularly enjoy reading fiction.”
22. “I find it hard to make new friends.” *
23. “I notice patterns in things all the time.”
24. “I would rather go to the theater than to a museum.”
25. “It does not upset me if my daily routine is disturbed.”
26. “I frequently find that I don’t know how to keep a conversation going.”
27. “I find it easy to “read between the lines” when someone is talking to me.”
28. “I usually concentrate more on the whole picture, rather than on the small details.”
29. “I am not very good at remembering phone numbers.”
30. “I don’t usually notice small changes in a situation or a person’s appearance.”
31. “I know how to tell if someone listening to me is getting bored.”
32. “I find it easy to do more than one thing at once.”
33. “When I talk on the phone, I’m not sure when it’s my turn to speak.”
34. “I enjoy doing things spontaneously.”
35. “I am often the last to understand the point of a joke.”
36. “I find it easy to work out what someone is thinking or feeling just by looking at their face.” *
37. “If there is an interruption, I can switch back to what I was doing very quickly.”
38. “I am good at social chitchat.”
39. “People often tell me that I keep going on and on about the same thing.”
40. “When I was young, I used to enjoy playing games involving pretending with other children.”
41. “I like to collect information about categories of things (e.g., types of cars, birds, trains, plants).”
42. “I find it difficult to imagine what it would be like to be someone else.”
43. “I like to carefully plan any activities I participate in.”
44. “I enjoy social occasions.” *
45. “I find it difficult to work out people’s intentions.” *
46. “New situations make me anxious.”
47. “I enjoy meeting new people.” *
48. “I am a good diplomat.” *
49. “I am not very good at remembering people’s date of birth.”
50. “I find it very easy to play games with children that involve pretending.”

(items with an asterisk indicate items belonging to the social skills subset)

**3. Subjective ratings of training questions**

1. ‘Did you find the tasks in the study tiring?’
2. ‘Did you find the tasks in the study interesting?’
3. ‘Did you find the tasks instructions in the study easy to follow?’
4. ‘Do you think that emotion recognition training would be useful as an intervention for people with autism?’.

**4. Additional pre-registered analyses**

We pre-registered additional analyses but these were more focused on developing our specific task and thus we have included details of these here.

We compared hits at baseline and in Session 4 for those who had a diagnosis of autism with those who identified as autistic (but did not indicate they have a diagnosis). For this, we compared data using plots and two-tailed independent means tests.

T-tests comparing hits for those with and without a diagnosis of autism revealed no meaningful differences at baseline (*p*=0.85) or in the Session 4 test (*p*=0.63) (also see Supplementary Figure S2).

We also plotted the data to explore training effects across Sessions 1 to 4, to examine how accuracy changes over the number of sessions, using the total number of correct responses for the first attempt in each training session.

Supplementary Figure S3 plots the total number of correct responses in the first attempt in each training session within the active group over time. It shows an increase in correct responses across time, suggesting experiencing more sessions leads to a greater number of correct responses.

**5. Skewness and kurtosis**

We assessed skewness and kurtosis for baseline and post-training (Session 4) total hits (on emotion recognition test). For baseline total hits skewness was -0.22 and kurtosis was -0.28. For Session 4 total hits skewness was -0.33 and kurtosis was -0.32. All values were within an acceptable range (Kim, 2013). Therefore, no transformations of these data were necessary.

**Supplementary Figure S1.** Histograms of the distribution of total hits as a proportion at baseline (a) and post-training (Session 4) (b).

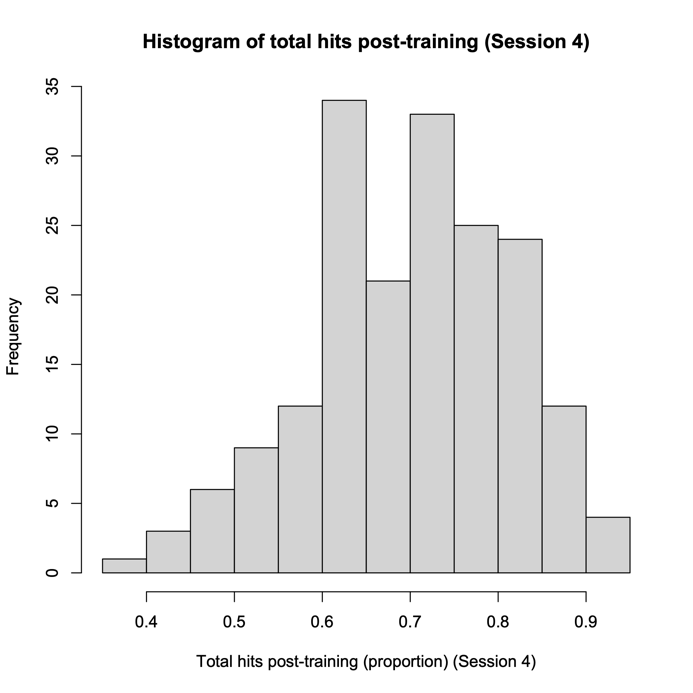

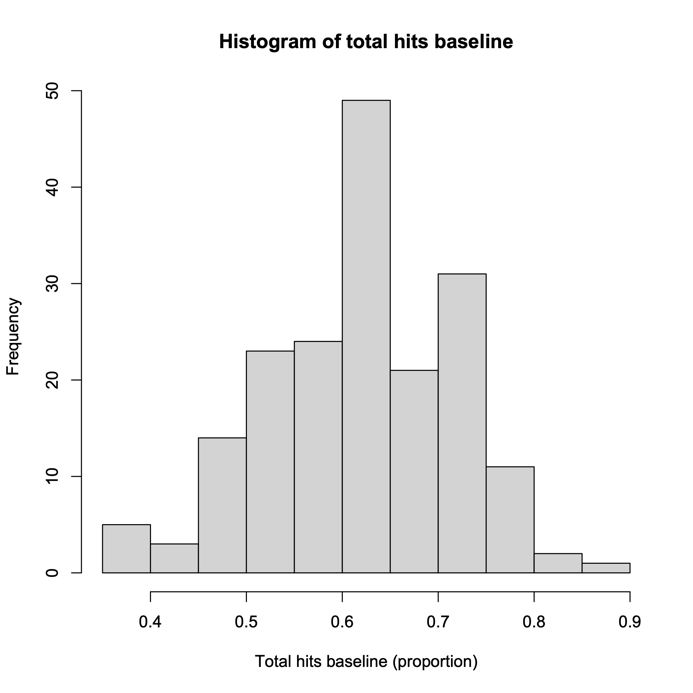

a)

b)

**Supplementary Figure S2.** Total hits at baseline and Session 4 for those with a diagnosis of autism compared to those without a formal diagnosis but who identified as autistic

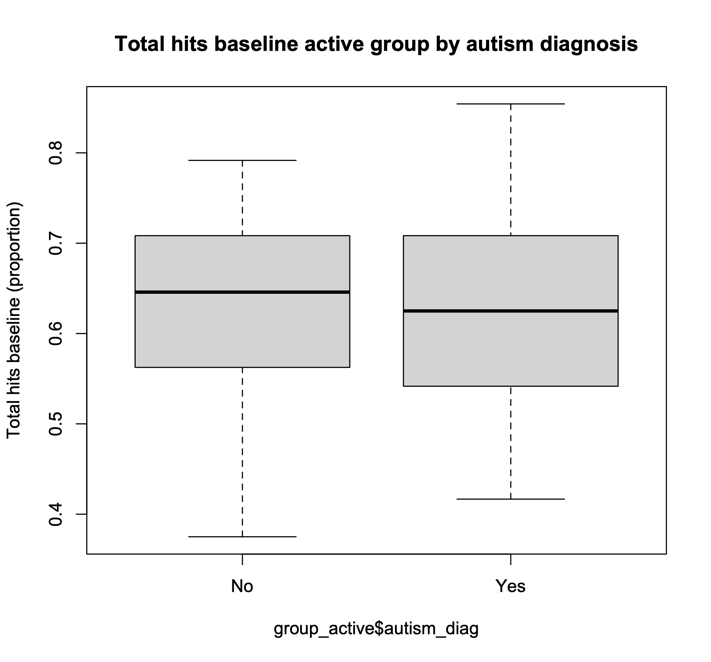

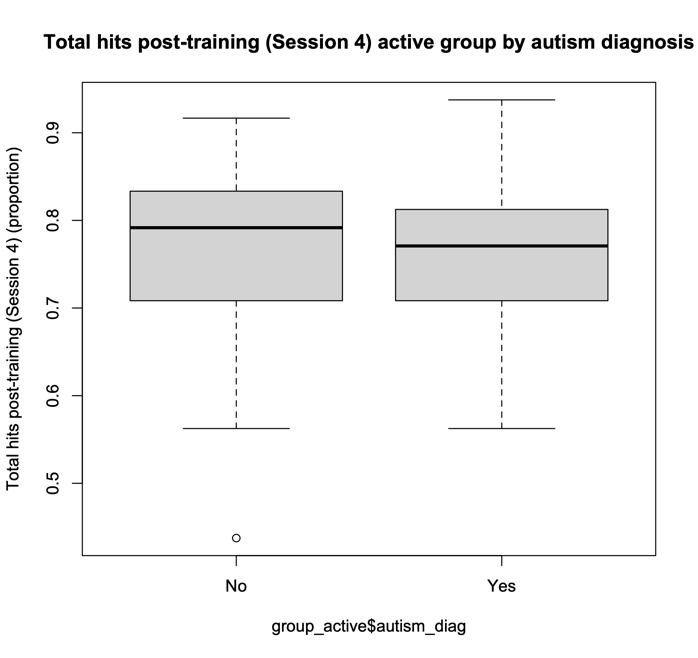

b)

a)

**Supplementary Figure S3.** Emotion recognition accuracy (mean number of total correct responses on the first attempt) over the 4 training sessions in the active group

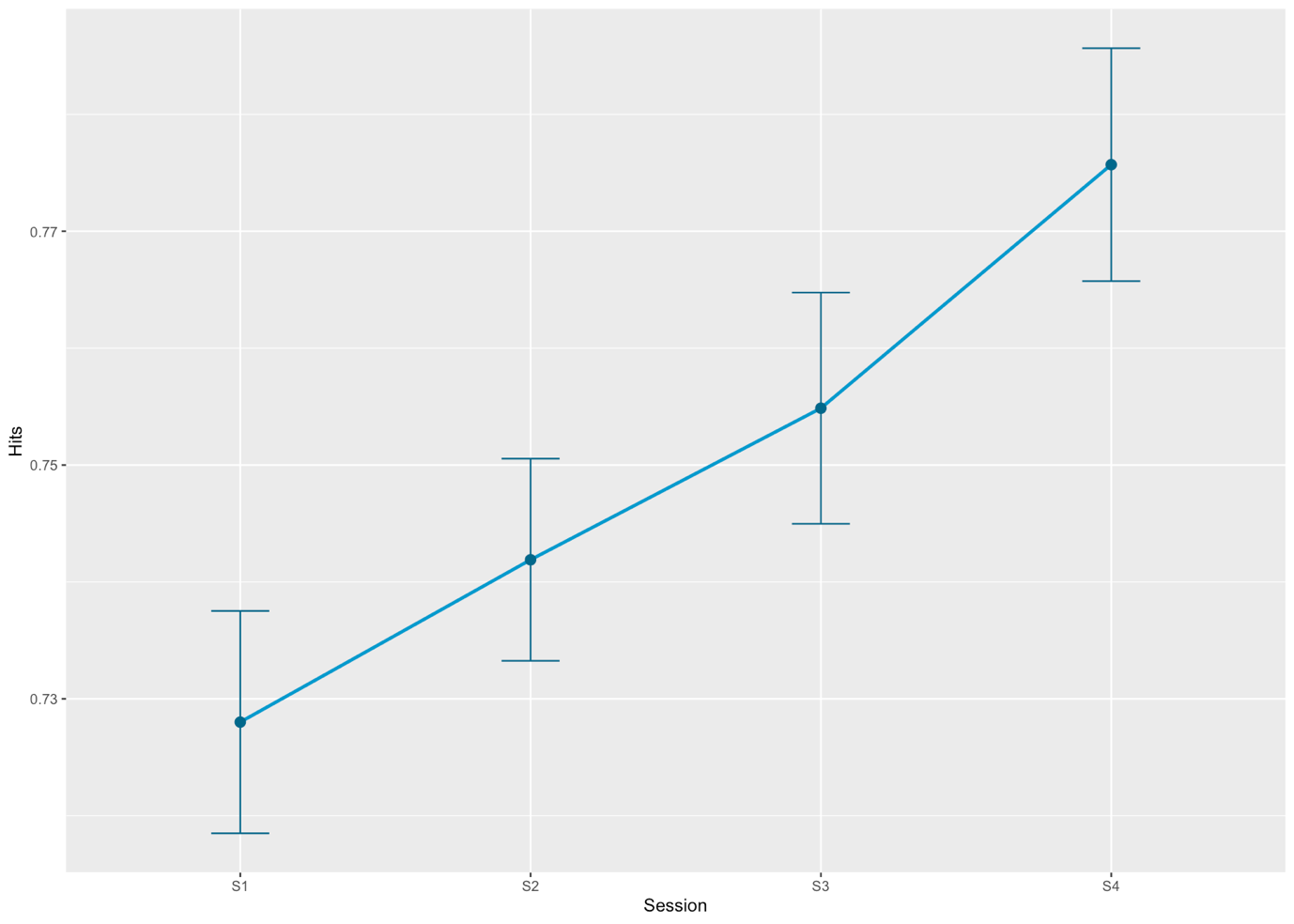

**Supplementary Table S1.** Fixed effects from LME model for total hits at baseline and Session 4, including an interaction between time and group (N=184).

|  | **Unadjusted** | | | | **Adjusted** | | | | **Adjusted for AQ50** | | | |
| --- | --- | --- | --- | --- | --- | --- | --- | --- | --- | --- | --- | --- |
|  | **Main effects model** | | **Interaction model (time*group)** | | **Main effects model** | | **Interaction model (time*group)** | | **Main effects model** | | **Interaction model (time*group)** | |
|  | ***b* (95% CI)** | ***p*** | ***b* (95% CI)** | ***p*** | ***b* (95% CI)** | ***p*** | ***b* (95% CI)** | ***p*** | ***b* (95% CI)** | ***p*** | ***b* (95% CI)** | ***p*** |
| **Intercept** | 0.59  (0.57 – 0.61) | 1.39x10^-167^ | 0.62 (0.60 – 0.64) | 2.65x10^-203^ | 0.64 (0.60 – 0.69) | 3.89x10^-69^ | 0.67 (0.63 – 0.72) | 7.80x10^-74^ | 0.70  (0.63 -0.77) | 3.93x10^-46^ | 0.73  (0.66- 0.80) | 1.36x10^-48^ |
| **Time [Session 4]** | 0.08 (0.06 – 0.10) | 7.26x10^-07^ | 0.02  (-0.01 – 0.05) | 0.13 | 0.08 (0.06 – 0.10) | 7.26x10^-02^ | 0.02  (-0.01 – 0.05) | 0.13 | 0.08  (0.06 – 0.10) | 7.26x10^-16^ | 0.02  (-0.01 – 0.05) | 0.13 |
| **Group [Active]** | 0.06 (0.04 – 0.09) | 5.60x10^-07^ | 0.01  (-0.02 – 0.04) | 0.68 | 0.06 (0.04 – 0.09) | 0.008 | 0.003  (-0.03 – 0.03) | 0.85 | 0.06  (0.04 – 0.09) | 8.76x10^-07^ | 0.01  (-0.03 – 0.04) | 0.69 |
| **Time [Session 4] * group [Active]** |  |  | 0.12 (0.08 – 0.16) | 4.00x10^-09^ |  |  | 0.12 (0.08 – 0.16) | 4.00x10^-09^ |  |  | 0.12  (0.08 – 0.16) | 4.00x10^-09^ |

*Results from the main effects and interaction LME models (with an interaction of time and group) are presented here, where the beta estimate indicates the amount by which the proportion of total hits is increased from baseline to Session 4. CI = confidence interval. LME = linear mixed effects.*

**Supplementary Table S2.** Fixed effects from LME model for total hits at baseline and Session 4, including an interaction between time and group, for analyses excluding those with a mental health diagnosis and technical difficulties, adjusted for age, gender and education.

|  | **Excluding those with a co-occurring mental health diagnosis (N=71)** | | | | **Excluding those who encountered technical difficulties (N=182)** | | | |
| --- | --- | --- | --- | --- | --- | --- | --- | --- |
|  | **Main effects model** | | **Interaction model (time*group)** | | **Main effects model** | | **Interaction model (time*group)** | |
|  | ***b* (95% CI)** | ***p*** | ***b* (95% CI)** | ***p*** | ***b* (95% CI)** | ***p*** | ***b* (95% CI)** | ***p*** |
| **Intercept** | 0.57 (0.48 – 0.66) | 7.69x10^-19^ | 0.60 (0.51 – 0.69) | 4.36x10^-20^ | 0.64 (0.60 -0.69) | 8.97x10^-69^ | 0.67 (0.63- 0.72) | 1.82x10^-73^ |
| **Time [Session 4]** | 0.08 (0.05 – 0.12) | 1.28x10^-07^ | 0.02 (-0.02 – 0.07) | 0.13 | 0.08 (0.06 – 0.10) | 5.52x10^-16^ | 0.02 (-0.01 – 0.05) | 0.13 |
| **Group [Active]** | 0.07 (0.02 – 0.11) | 0.003 | 0.01 (-0.04 – 0.07) | 0.65 | 0.06 (0.04 – 0.09) | 2.27x10^-06^ | 0.003 (-0.03 – 0.03) | 0.87 |
| **Time [Session 4] * group [Active]** |  |  | 0.11 (0.05 – 0.17) | 0.001 |  |  | 0.12 (0.08 – 0.16) | 2.08x10^-09^ |

*Results from the main effects and interaction LME models (with an interaction of time and group) are presented here, where the beta estimate indicates the amount by which the proportion of total hits is increased from baseline to Session 4, when excluding participants with co-occurring mental health diagnoses and who encountered technical difficulties. CI = confidence interval. LME = linear mixed effects.*

**Supplementary Table S3.** Fixed effects from LME model for total hits at baseline and generalisability test in Session 4, including an interaction between time and group, adjusted for age, gender and education (N=184).

|  | **Main effects model** | | **Interaction model (time*group)** | |
| --- | --- | --- | --- | --- |
|  | ***b* (95% CI)** | ***p*** | ***b* (95% CI)** | ***p*** |
| **Intercept** | 0.66 (0.61 – 0.70) | 5.27x10^-73^ | 0.67 (0.63 – 0.72) | 2.18x10^-76^ |
| **Time [generalisability Session 4]** | 0.08 (0.06 – 0.10) | 1.25x10^-14^ | 0.05 (0.02 – 0.08) | 0.0006 |
| **Group [Active]** | 0.03 (0.01 – 0.05) | 0.01 | 0.003 (-0.03 – 0.03) | 0.87 |
| **Time [generalisability Session 4] * group [Active]** |  |  | 0.06 (0.02 – 0.09) | 0.005 |

*Results from the main effects and interaction LME models (with an interaction of time and group) are presented here, where the beta estimate indicates the amount by which the proportion of total hits is increased from baseline to the generalisability test. CI = confidence interval. LME = linear mixed effects.*

**Supplementary Table S4.** Fixed effects from LME model for total hits at baseline and Session 5, including an interaction between time and group, adjusted for age, gender and education and additional analyses excluding those excluding those with a mental health diagnosis or with technical difficulties.

|  | **Adjusted analysis (N=169)** | | | | **Excluding those who encountered technical difficulties (N=160)** | | | | **Excluding those with a co-occurring mental health diagnosis (N=65)** | | | |
| --- | --- | --- | --- | --- | --- | --- | --- | --- | --- | --- | --- | --- |
|  | **Main effects model** | | **Interaction model (time*group)** | | **Main effects model** | | **Interaction model (time*group)** | | **Main effects model** | | **Interaction model (time*group)** | |
|  | ***b* (95% CI)** | ***p*** | ***b* (95% CI)** | ***p*** | ***b* (95% CI)** | ***p*** | ***b* (95% CI)** | ***p*** | ***b* (95% CI)** | ***p*** | ***b* (95% CI)** | ***p*** |
| **Intercept** | 0.65 (0.60 – 0.70) | 7.07x10^-62^ | 0.67 (0.62 – 0.72) | 8.84x10^-65^ | 0.65  (0.60-0.70) | 3.46x10^-58^ | 0.67  (0.62 – 0.72) | 6.53x10^-61^ | 0.57  (0.48 - 0.67) | 1.79x10^-17^ | 0.60  (0.50 – 0.70) | 1.78x10^-18^ |
| **Time [Session 5]** | 0.08 (0.06 – 0.10) | 4.89x10^-14^ | 0.04 (0.01 – 0.07) | 0.003 | 0.08  (0.06-0.10 | 8.04x10^-13^ | 0.04  (0.01 – 0.07) | 0.007 | 0.08  (0.05 - 0.11 | 8.02x10^-07^ | 0.03  (-0.02 – 0.08) | 0.24 |
| **Group [Active]** | 0.04 (0.01 – 0.07) | 0.004 | 0.01  (-0.03 – 0.04) | 0.75 | 0.03  (0.01- 0.06) | 0.02 | -0.0004  (-0.04 – 0.03) | 0.98 | 0.06  (0.02 - 0.11) | 0.006 | 0.02  (-0.04 – 0.07) | 0.54 |
| **Time [Session 5] * group [Active]** |  |  | 0.07 (0.03 – 0.11) | 0.001 |  |  | 0.07  (0.03 – 0.11) | 0.001 |  |  | 0.09  (0.03 – 0.15) | 0.005 |

*Results from the main effects and interaction LME models (with an interaction of time and group) are presented here, where the beta estimate indicates the amount by which the proportion of total hits is increased from baseline to Session 5. CI = confidence interval. LME = linear mixed effects.*

**Supplementary Table S5.** Results for the interaction between time (Session 4) and group (active) from LME models for emotion specific sensitivity scores (N=184).

|  | **Angry** | | | | **Happy** | | | | **Sad** | | | | **Scared** | | | | **Surprised** | | | | **Disgust** | | | |
| --- | --- | --- | --- | --- | --- | --- | --- | --- | --- | --- | --- | --- | --- | --- | --- | --- | --- | --- | --- | --- | --- | --- | --- | --- |
|  | **Main effects model** | | **Interaction model (time*group)** | | **Main effects model** | | **Interaction model (time*group)** | | **Main effects model** | | **Interaction model (time*group)** | | **Main effects model** | | **Interaction model (time*group)** | | **Main effects model** | | **Interaction model (time*group)** | | **Main effects model** | | **Interaction model (time*group)** | |
|  | ***b* (95% CI)** | ***p*** | ***b* (95% CI)** | ***p*** | ***b* (95% CI)** | ***p*** | ***b* (95% CI)** | ***p*** | ***b* (95% CI)** | ***p*** | ***b* (95% CI)** | ***p*** | ***b* (95% CI)** | ***p*** | ***b* (95% CI)** | ***p*** | ***b* (95% CI)** | ***p*** | ***b* (95% CI)** | ***p*** | ***b* (95% CI)** | ***p*** | ***b* (95% CI)** | ***p*** |
| Intercept | 0.93  (0.90 - 0.96) | 1.69x10^-114^ | 0.94 (0.91 - 0.98) | 1.40x10^-117^ | 0.88 (0.86 - 0.91) | 2.72x10^-134^ | 0.90 (0.87 - 0.93) | 1.35x10^-139^ | 0.92 (0.90 - 0.94) | 1.09x10^-164^ | 0.93 (0.91 - 0.95) | 6.97x10^-174^ | 0.73 (0.65 - 0.82) | 1.99x10^-38^ | 0.77 (0.68 - 0.85) | 2.54x10^-40^ | 0.86 (0.83 - 0.89) | 2.05x10^-121^ | 0.86 (0.83 - 0.89) | 2.87x10^-127^ | 0.93 (0.88 - 0.98) | 1.54x10^-82^ | 0.94 (0.89 - 0.99) | 1.07x10^-84^ |
| Time [Session 4] | 0.03  (0.02 - 0.04) | 1.79x10^-07^ | 0.005 (-0.01 - 0.02) | 0.53 | 0.03 (0.02 - 0.04) | 1.93x10^-06^ | -0.01 (-0.02 - 0.01) | 0.38 | 0.02 (0.01 - 0.03) | 1.23x10^-04^ | 0.0003 (-0.01 - 0.01) | 0.96 | 0.11 (0.08 - 0.14) | 2.17x10^-10^ | 0.05 (0.003 - 0.09) | 0.04 | 0.02 (0.004 - 0.03) | 0.01 | 0.02 (0.002 - 0.04) | 0.04 | 0.03 (0.01 - 0.04) | 0.003 | 0.02 (-0.01 - 0.04) | 0.22 |
| Group [Active] | 0.04  (0.02 - 0.05) | 2.47x10^-04^ | 0.01 (-0.01 - 0.03) | 0.27 | 0.02 (0.01 - 0.04) | 0.004 | -0.01 (-0.03 - 0.003) | 0.12 | 0.02 (0.01 - 0.03) | 0.002 | -0.0002 (-0.01 - 0.01) | 0.98 | 0.05 (0.0007 - 0.09) | 0.05 | -0.01 (-0.07 - 0.04) | 0.60 | 0.02 (0.004 - 0.04) | 0.02 | 0.02 (0.002 - 0.05) | 0.03 | 0.03 (0.002 - 0.06) | 0.04 | 0.02 (-0.01 - 0.05) | 0.27 |
| Time [Session 4] * group [Active] |  |  | 0.05 (0.03 - 0.07) | 4.90x10^-06^ |  |  | 0.07 (0.05 - 0.09) | 3.77x10^-10^ |  |  | 0.03 (0.02 - 0.05) | 1.24x10^-04^ |  |  | 0.12 (0.06 - 0.18) | 8.60x10^-05^ |  |  | -0.01 (-0.04 - 0.02) | 0.60 |  |  | 0.02 (-0.01 - 0.06) | 0.23 |

*Results from the adjusted (for age, gender and education level) main effects and interaction LME models for sensitivity scores (with an interaction of time and group) are presented here, where the beta estimate indicates the amount by which the sensitivity score is increased. CI = confidence interval. LME = linear mixed effects*

**Supplementary Table S6.** Results for the interaction between time (Session 4) and group (active) from LME models for emotion specific hits (N=184).

|  | **Angry** | | | | **Happy** | | | | **Sad** | | | | **Scared** | | | | **Surprised** | | | | **Disgust** | | | |
| --- | --- | --- | --- | --- | --- | --- | --- | --- | --- | --- | --- | --- | --- | --- | --- | --- | --- | --- | --- | --- | --- | --- | --- | --- |
|  | **Main effects model** | | **Interaction model (time*group)** | | **Main effects model** | | **Interaction model (time*group)** | | **Main effects model** | | **Interaction model (time*group)** | | **Main effects model** | | **Interaction model (time*group)** | | **Main effects model** | | **Interaction model (time*group)** | | **Main effects model** | | **Interaction model (time*group)** | |
|  | ***b* (95% CI)** | ***p*** | ***b* (95% CI)** | ***p*** | ***b* (95% CI)** | ***p*** | ***b* (95% CI)** | ***p*** | ***b* (95% CI)** | ***p*** | ***b* (95% CI)** | ***p*** | ***b* (95% CI)** | ***p*** | ***b* (95% CI)** | ***p*** | ***b* (95% CI)** | ***p*** | ***b* (95% CI)** | ***p*** | ***b* (95% CI)** | ***p*** | ***b* (95% CI)** | ***p*** |
| Intercept | 0.65 (0.58 - 0.73) | 7.44x10^-38^ | 0.70 (0.62 - 0.78) | 5.38x10^-41^ | 0.56 (0.46 - 0.66) | 2.06x10^-22^ | 0.63 (0.53 - 0.73) | 5.31x10^-26^ | 0.71 (0.65 - 0.77) | 1.50x10^-54^ | 0.73 (0.67 - 0.80) | 4.50x10^-57^ | 0.33 (0.20 - 0.46) | 1.47x10^-06^ | 0.38 (0.25 - 0.51) | 4.41x10^-08^ | 0.74 (0.68 - 0.81) | 1.57x10^-53^ | 0.73 (0.66 - 0.79) | 1.44x10^-52^ | 0.87 (0.76 - 0.97) | 2.44x10^-39^ | 0.88 (0.78 - 0.98) | 4.83x10^-40^ |
| Time [Session 4] | 0.09 (0.06 - 0.12) | 3.22x10^-08^ | 0.004 (-0.04 - 0.04) | 0.84 | 0.13 (0.09 - 0.17) | 5.98x10^-10^ | 0.004 (-0.05 - 0.05) | 0.87 | 0.03 (0.01 - 0.06) | 0.02 | -0.01 (-0.05 - 0.02) | 0.47 | 0.20 (0.15 - 0.24) | 1.87x10^-16^ | 0.09 (0.03 - 0.15) | 0.002 | -0.01 (-0.04 - 0.02) | 0.42 | 0.02  (-0.02 - 0.06) | 0.33 | 0.05 (0.01 - 0.08) | 0.02 | 0.02  (-0.03 - 0.08) | 0.45 |
| Group [Active] | 0.11 (0.07 - 0.15) | 9.13x10^-07^ | 0.03 (-0.02 - 0.08) | 0.29 | 0.05 (-0.01 - 0.10) | 0.08 | -0.08 (-0.14 -0.01) | 0.02 | 0.06 (0.03 - 0.09) | 0.001 | 0.01 (-0.03 - 0.06) | 0.54 | 0.08 (0.01 - 0.16) | 0.02 | -0.02 (-0.10 - 0.06) | 0.67 | 0.02 (-0.02 - 0.05) | 0.41 | 0.05 (0.001 - 0.09) | 0.05 | 0.05 (-0.01 - 0.10) | 0.09 | 0.02  (-0.04 - 0.09) | 0.49 |
| Time [Session 4] * group [Active] |  |  | 0.17 (0.11 - 0.22) | 2.13x10^-08^ |  |  | 0.25 (0.18 - 0.32) | 6.19x10^-11^ |  |  | 0.09 (0.04 - 0.15) | 0.001 |  |  | 0.20 (0.12 - 0.28) | 1.24x10^-06^ |  |  | -0.06  (-0.12 -0.01) | 0.03 |  |  | 0.05  (-0.03 - 0.13) | 0.20 |

*Results from the adjusted (for age, gender and education level) main effects and interaction LME models for emotion specific hits (with an interaction of time and group) are presented here, where the beta estimate indicates the amount by which the proportion of total hits is increased. CI = confidence interval. LME = linear mixed effects*

**Supplementary Table S7.** Results for the interaction between time (Session 4) and group (active) from LME models for emotion specific false alarms (N=184).

|  | **Angry** | | | | **Happy** | | | | **Sad** | | | | **Scared** | | | | **Surprised** | | | | **Disgust** | | | |
| --- | --- | --- | --- | --- | --- | --- | --- | --- | --- | --- | --- | --- | --- | --- | --- | --- | --- | --- | --- | --- | --- | --- | --- | --- |
|  | **Main effects model** | | **Interaction model (time*group)** | | **Main effects model** | | **Interaction model (time*group)** | | **Main effects model** | | **Interaction model (time*group)** | | **Main effects model** | | **Interaction model (time*group)** | | **Main effects model** | | **Interaction model (time*group)** | | **Main effects model** | | **Interaction model (time*group)** | |
|  | ***b* (95% CI)** | ***p*** | ***b* (95% CI)** | ***p*** | ***b* (95% CI)** | ***p*** | ***b* (95% CI)** | ***p*** | ***b* (95% CI)** | ***p*** | ***b* (95% CI)** | ***p*** | ***b* (95% CI)** | ***p*** | ***b* (95% CI)** | ***p*** | ***b* (95% CI)** | ***p*** | ***b* (95% CI)** | ***p*** | ***b* (95% CI)** | ***p*** | ***b* (95% CI)** | ***p*** |
| Intercept | 0.02 (0.004 - 0.04) | 0.02 | 0.02  (-0.002 - 0.04) | 0.04 | 0.03 (0.01 - 0.06) | 0.01 | 0.04 (0.01 - 0.066) | 0.009 | 0.02  (-0.01 - 0.05) | 0.16 | 0.02  (-0.01 - 0.05) | 0.24 | 0.06 (0.04 - 0.08) | 1.12x10^-07^ | 0.05  (0.03 - 0.08) | 1.86x10^-06^ | 0.16 (0.13 - 0.20) | 2.86x10^-19^ | 0.15 (0.12 - 0.18) | 1.75x10^-16^ | 0.12 (0.10 - 0.15) | 4.06x10^-15^ | 0.11  (0.08 - 0.14) | 1.32x10^-12^ |
| Time [Session 4] | -0.002  (-0.01 - 0.005) | 0.51 | 0.002  (-0.01 - 0.01) | 0.66 | 0.01 (0.003 - 0.02) | 0.008 | 0.01  (-0.005 - 0.02) | 0.21 | -0.02 (-0.03 -0.01) | 9.43x10^-05^ | -0.01  (-0.03 - 0.0009) | 0.07 | -0.01  (-0.02 -0.005) | 0.002 | -0.0008 (-0.01 - 0.01) | 0.88 | -0.05  (-0.07 -0.43) | 3.43x10^-15^ | -0.02  (-0.04 -0.01) | 0.005 | -0.02  (-0.03 -0.01) | 0.001 | 0.01  (-0.01 - 0.02) | 0.50 |
| Group [Active] | -0.002  (-0.01 - 0.01) | 0.74 | 0.003  (-0.01 - 0.02) | 0.68 | -0.01  (-0.03 - 0.001) | 0.08 | -0.02 (-0.03 - 0.0002) | 0.06 | 0.002 (-0.02 - 0.02) | 0.81 | 0.01  (-0.01 - 0.03) | 0.38 | -0.01  (-0.02 - 0.0005) | 0.07 | 0.0002  (-0.01 - 0.01) | 0.97 | -0.03  (-0.05 -0.01) | 3.96x10^-04^ | -0.003  (-0.02 - 0.02) | 0.75 | -0.02  (-0.04 -0.004) | 0.01 | 0.005  (-0.01 - 0.02) | 0.64 |
| Time [Session 4] * group [Active] |  |  | -0.01  (-0.02 - 0.005) | 0.20 |  |  | 0.01  (-0.01 - 0.03) | 0.40 |  |  | -0.01  (-0.03 - 0.01) | 0.18 |  |  | -0.02  (-0.04 -0.01) | 0.004 |  |  | -0.06  (-0.08 - 0.03) | 1.61x10^-06^ |  |  | -0.05  (-0.07 - 0.03) | 1.97x10^-05^ |

*Results from the adjusted (for age, gender and education level) main effects and interaction LME models for false alarms (with an interaction of time and group) are presented here, where the beta estimate indicates the amount by which the proportion of false alarms score is increased. CI = confidence interval. LME = linear mixed effects*

**Supplementary Table S8.** Fixed effects from LME model for social skills (as measured using a subset of the AQ-50) at baseline and Session 4, and baseline and Session 5, including an interaction between time and group.

|  | **Session 4 (N=184)** | | | | | | | | **Session 5 (N=169)** | | | | | | | |
| --- | --- | --- | --- | --- | --- | --- | --- | --- | --- | --- | --- | --- | --- | --- | --- | --- |
|  | **Unadjusted** | | | | **Adjusted** | | | | **Unadjusted** | | | | **Adjusted** | | | |
|  | **Main effects model** | | **Interaction model (time*group)** | | **Main effects model** | | **Interaction model (time*group)** | | **Main effects model** | | **Interaction model (time*group)** | | **Main effects model** | | **Interaction model (time*group)** | |
|  | ***b* (95% CI)** | ***p*** | ***b* (95% CI)** | ***p*** | ***b* (95% CI)** | ***p*** | ***b* (95% CI)** | ***p*** | ***b* (95% CI)** | ***p*** | ***b* (95% CI)** | ***p*** | ***b* (95% CI)** | ***p*** | ***b* (95% CI)** | ***p*** |
| **Intercept** | 6.59  (6.20 - 6.98) | 5.33x10^-08^ | 6.60  (6.20 - 7.00) | 2.17x10^-86^ | 6.33  (5.38 - 7.29) | 8.90x10^-28^ | 6.34  (5.38 - 7.30) | 8.59x10^-28^ | 6.70  (6.31 - 7.10) | 1.32x10^-79^ | 6.78  (6.38 - 7.19) | 1.55x10^-82^ | 6.61  (5.65 - 7.57) | 1.77x10^-28^ | 6.69  (5.73 - 7.65) | 6.41x10^-29^ |
| **Time [Session 4/5]** | -0.15  (-0.32 - 0.03) | 0.10 | -0.17  (-0.42 - 0.08) | 0.19 | -0.15  (-0.32 - 0.03) | 0.10 | -0.17  (-0.42 - 0.08) | 0.19 | -0.23  (-0.40 -0.06) | 0.009 | -0.39  (-0.63 -0.15) | 0.001 | -0.23  (-0.40 -0.06) | 0.009 | -0.39  (-0.63 -0.15) | 0.002 |
| **Group [Active]** | 0.38  (-0.15 - 0.91) | 0.16 | 0.36  (-0.20 - 0.91) | 0.21 | 0.35  (-0.18 - 0.88) | 0.19 | 0.33  (-0.22 - 0.89) | 0.24 | 0.38  (-0.16 - 0.92) | 0.17 | 0.22  (-0.35 - 0.78) | 0.45 | 0.40  (-0.14 - 0.94) | 0.15 | 0.24  (-0.33 - 0.81) | 0.41 |
| **Time [Session 4/ 5] * group [Active]** |  |  | 0.04  (-0.31 - 0.39) | 0.83 |  |  | 0.04  (-0.31 - 0.39) | 0.83 |  |  | 0.32  (-0.01 - 0.66) | 0.06 |  |  | 0.32  (-0.01 - 0.66) | 0.06 |

*Results from the main effects and interaction LME models (with an interaction of time and group) are presented here, where the beta estimate indicates the amount by which the social skills score is increased from baseline to Session 4 or Session 5. CI = confidence interval. LME = linear mixed effects.*

**Supplementary Table S9.** Results from t-tests of group differences in subjective ratings, other social skills, and training experience questions.

|  | **Active group**  **mean (SD)** | **Sham group**  **mean (SD)** | ***p*** |
| --- | --- | --- | --- |
| ***Subjective ratings (N=184)*** | | | |
| **Tiring** | 48 (27) | 45 (29) | 0.51 |
| **Interesting** | 62 (27) | 52 (25) | 0.01 |
| **Easy to follow** | 96 (9) | 90 (15) | 0.002 |
| ***Social skills session 4 (N=184)*** | | | |
| **Frequency** | 20 (26) | 17 (23) | 0.40 |
| **Improved ER** | 40 (27) | 26 (23) | 0.0002 |
| ***Social skills session 5 (N=168)*** | | | |
| **Frequency** | 38 (30) | 28 (28) | 0.03 |
| **Improved ER** | 47 (24) | 33 (22) | 9.19x10^-05^ |
| ***Training experience (N=184)*** | | | |
| **Is training useful?** | 69 (23) | 68 (21) | 0.88 |

*The mean and standard deviations (SD) for each item per group are presented here along with the p-value from t-tests conducted. We are comparing these ratings across groups as the tasks completed were different and we might expect there to be differences in how participants found the tasks. We also wanted to see whether there was any indication of differences in social skills outcomes for those that received the emotion recognition training compared to those that did not. ER = emotion recognition.*
